# Psychometric properties of the Perceived Stress Scale-4 (PSS-4) in Colombians with university education

**DOI:** 10.1101/2023.09.05.23295052

**Authors:** Juan P. Sanabria-Mazo, Andrés Gómez-Acosta, Julio Annicchiarico-Lobo, Juan V. Luciano, Antoni Sanz

## Abstract

The Perceived Stress Scale-4 (PSS-4) is an ultra-brief self-report measure to assess perceived psychological stress. This study evaluated the psychometric properties of the PSS-4 in Colombia. A total of 1911 adult participants with university education completed the PSS-4. Characteristics of the items and the subscales were explored. Dimensionality was assessed using confirmatory factor analysis (CFA), incorporating an examination of invariance (configural, metric, scalar, and strict) across socio-demographic characteristics. Construct validity (convergent and discriminant), reliability indices, know-groups, and predictive validity were also computed. The CFA showed adequate fit indices for the two-factor model (hopelessness and coping), being invariant across gender, income level, age, and work status. Internal consistency was adequate for the PSS-4 (*α* = .73, *ω* = .72, *λ*^2^ = .74). Significant positive correlations of the PSS-4 were identified with depression (*r* = .59) and anxiety (*r* = .55), as well as significant negative with post-traumatic growth (*r* = –.37) and resilience (*r* = –.47). The PSS-4 showed adequate capacity to predict potential depressive and anxiety symptoms, as well as protective factors such as resilience and posttraumatic growth. Higher scores on the PSS-4 were observed among young people, as well as among people with lower incomes and those unemployed. These findings suggest that the PSS-4 can be a reliable and valid tool for assessing psychological stress in Colombians with university education.

## Introduction

Stress is a common experience in industrialized societies, and it’s associated with a high public health and societal impact (Cohen et al., 2007; McEwen, 2017; Xiong et al., 2020). Over time, it has been conceptualized mainly from three complimentary approaches: (1) psychological, based on people’s subjective appraisals (Lazarus & Folkman, 1984); (2) biological, related to physiological responses to stress (McEwen, 2007); and (3) environmental, associated to life events (Monroe & Simons, 1991). From a psychological transactional approach, stress has been defined as the reaction that occurs when people perceive a discrepancy between their resources and/or their capacity to respond to an event, stimulus, or stressor (Lazarus & Folkman, 1984).

For many years, psychological stress has been linked to negative health status, due to increased allostatic load through psychophysiological reactivity (Sanz et al., 2007), epigenetic mechanisms (Gómez-Acosta & Londoño, 2021; Lee et al., 2023; Mauss & Jarkzoc, 2021; Zhu et al., 2023), and poor mental health outcomes (Feng et al., 2023). In fact, there is evidence that chronic psychological stress is associated with 75-90% of health conditions (Liu et al., 2017). The early detection and prevention of psychological stress symptoms, thus avoiding their chronification, is a challenge of increasing interest for clinicians and researchers worldwide (Gómez-Acosta & Castro, 2020; Xiong et al., 2020).

A progressive increase in psychological stress had been documented in all populations in the last decades (Enticott et al., 2022). Already during the coronavirus disease 2019 (COVID-19) pandemic, an acute increase in psychological stress levels was documented in 57 countries, particularly in women, older adults, and people with low educational level (Adamson et al., 2020). In the post-pandemic era, an increase in the prevalence of mental health disorders have been widely reported, often related to stressors not directly controllable by the people, such as natural disasters (World Economic Forum, 2023) and unfavorable political decisions (American Psychological Association [APA], 2023). These events have especially impacted populations with high social and economic inequality, such as migrants (World Economic Forum, 2023), workers from different areas (Stringer, 2023a), and university staff (Stringer, 2023b).

A timely assessment with an adequate approach constitutes an invaluable contribution to a reduction of psychological stress as a risk factor of suicide, as referred to in the extended Comprehensive Mental Health Action Plan 2013–2030 (World Health Organization [WHO], 2021), and for buffering the development of emotional disorders, such as depression, anxiety disorders, and post-traumatic stress disorder (Cuijpers et al., 2008; Feng et al., 2023; Xiong et al., 2020). In this context, it is important to have reliable, valid, and brief screening tools for measuring psychological stress (Golberg et al., 2017). Particularly, ultra-brief screening tools could help reduce misdiagnosis, optimize healthcare system resources, and improve clinical outcomes (Castro-Rodríguez et al., 2015; Sanabria-Mazo et al., 2023).

The Perceived Stress Scale (PSS) is a widely used tool for assessing perceived psychological stress. The psychometric properties of PSS-14 and PSS-10 versions have been extensively explored in recent years in clinical and non-clinical populations (Golden-Kreutz et al., 2004; Lee et al., 2015; Maroufizadeh et al., 2018; Schneider et al., 2020), obtaining results demonstrating their validity and reliability. It has also been translated into Spanish (Remor, 2006), with evaluations of cross-cultural factorial invariance (Perera et al., 2017) and adaptations with samples of population with university education, specifically in the Spanish-speaking context in Colombia (Campo-Arias et al., 2009), Mexico (González-Ramírez et al., 2013), and Peru (Boluarte-Carbajal et al., 2023; Guzmán-Yacaman & Reyes-Bossio, 2018). The PSS-4, an ultra-brief version of four items, has also been tested in England (Warttig et al., 2013), the United Kingdom, France, and Spain (Vallejo et al., 2018), and Ecuador (Ruisoto et al., 2020), with adequate psychometrics properties in the one-factor (perceived psychological stress) and two-factor models (hopelessness and coping).

As far as it is known, the psychometric properties have not been examined in an ultra-brief version for the Colombian population, a highly distressed population in the last decades due to armed in conflicts and trauma exposure (Chaskel et al., 2015; Cuartas-Ricaurte et al., 2019; Gómez-Restrepo et al., 2016). Thus, the main objective of this study was to evaluate the psychometric properties of the PSS-4 in Colombia. The specific objectives and hypotheses explored in this research were:

1. Firstly, to examine the goodness-of-fit of three models: (1) One-factor model (i.e., psychological stress) with the four items loading on one latent factor, (2) a unidimensional model (i.e., psychological stress) with method effects (correlated error terms on the negative-phrased items), and (3) a two-factor model with two items in each factor (i.e., hopelessness and coping). Considering the evidence reported in previous studies (Boluarte-Carbajal et al., 2023; Ruisoto et al., 2020), it was speculated that the two-factor model would have a significantly better fit than the one-factor model (hypothesis 1).
2. Secondly, to investigate invariance (i.e., configural, metric, scalar, and strict) of the best-fitting model across socio-demographic characteristics (i.e., gender, income level, age range, and work status). To the best of this research team’s knowledge, this is the first study to provide evidence for invariance in PSS-4. As in previous validations of PSS-10 (Barbosa-Leiker et al., 2013; Lee, 2022; Liu et al., 2020; Reis et al., 2019), it was expected that the scores were invariant across gender (hypothesis 2).
3. Thirdly, to explore the reliability of the PSS-4 through different reliability indexes (i.e., Cronbach’s *α*, McDonald’s *ω*, and Guttman’s *λ*^2^). Being consistent with findings reported in previous studies (Vallejo et al., 2018; Warttig et al., 2013; Ruisoto et al., 2020), the PSS-4 was expected to reliably measure psychological stress (one-factor model), as well as hopelessness and coping (two-factor model) beyond the reliability index examined (hypothesis 3).
4. Fourthly, to analyze the construct validity (convergent and discriminant). In congruence with results reported in other studies (Vallejo et al., 2018; Warttig et al., 2013; Ruisoto et al., 2020), the PSS-4 scores are expected to be positively correlated with anxiety and depression and negatively with post-traumatic growth and resilience (hypothesis 4).
5. Fifthly, to evaluate the predictive validity of the PSS-4 on a set of psychological variables (resilience, posttraumatic growth, anxiety, and depression). As in previous studies (APA, 2023; Feng et al., 2023; Finstad et al., 2021; Gómez-Acosta et al., 2023), PSS-4 scores were expected to significantly predict resilience, posttraumatic growth scores, anxiety, and depression (hypothesis 5).
6. Sixthly, to test the relationship between the PPS-4 scores with socio-demographic characteristics of this sample. Based on the results from previous psychometric studies (Vallejo et al., 2018; Warttig et al., 2013; Ruisoto et al., 2020) and from a known-groups validity approach, it was expected that females, older individuals, those with lower incomes, or those unemployed would exhibit higher PSS-4 scores (hypothesis 6).

## Method

### Study design

This is a cross-sectional online psychometric study (Ato et al., 2013). Data analyses of the PSS-4 were performed using the dataset from the PSY-COVID study in Colombia (Sanz et al., 2023). More detailed information about this project is provided elsewhere (Sanabria-Mazo et al., 2021).

### Participants

This study included adult participants (≥ 18 years) with university education residing in Colombia. A total of 1911 adults (*M*_age_ = 38.40; *SD*_age_ = 11.82; range = 19-82) participated in the study. As shown in Table 1, most participants were female (79%), adults between 30-44 years old (41%), with medium income levels (65%), and formal workers (56%).

**Table 1.** Socio-demographic characteristics of the sample.

| **Variables, *n* (%)** | **Sample (*n* = 1911)** |
| --- | --- |
| *Gender* |  |
| Male | 402 (21) |
| Female | 1509 (79) |
| *Age group* |  |
| 18-29 years | 561 (29.4) |
| 30-44 years | 785 (41) |
| ≥ 45 years | 565 (29.6) |
| *Income level* |  |
| Low | 490 (25.6) |
| Medium | 1250 (65.4) |
| High | 171 (8.9) |
| *Work status* |  |
| Formal workers (with a contract) | 1062 (55.6) |
| Informal workers (without a contract) | 338 (17.7) |
| Unemployed | 329 (17.2) |
| Others | 182 (9.5) |

*Note.* *n* = frequency, % = percentage.

### Procedure

An anonymous survey was carried out using Google Forms® through non-probabilistic sampling. This survey was disseminated through social networks, media, and institutional contacts. The data collection for responses was available from December 1^st^ (2021) to April 30^th^ (2022). Participants were not provided with any economic incentives to complete the survey, which lasted 4 around minutes. This research was approved by the Animal and Human Experimentation Ethics Committee of the Autonomous University of Barcelona (CEEAH-5197) and was conducted according to the principles reported in the Helsinki Declaration.

### Instruments

*Sociodemographic Questionnaire.* This instrument was used to identifies the gender (male and female), age, income level (low, medium, and high), and work status (informal workers, formal workers, unemployed, and others).

*Perceived Stress Scale (PSS-4).* This instrument assesses perceived stress and/or the degree to which people evaluate their lives as stressful, especially regarding how uncontrollable and overloading relevant aspects of life are perceived (Cohen et al., 1983). There are three versions of the PSS (PSS-4, PSS-10, and PSS-14), although in this study only the ultra-brief version is explored. This version contains 4 items with a 4-point Likert response format, where 0 corresponds to “never” and 3 to “almost every day”. In the present work, the total score of the PSS-4 ranged from 0 to 12 (instead of 0-16), with higher scores indicating higher perceived stress. In the context of the development and analysis of ultra-brief versions of screening instruments, a 4-point response scale was incorporated as an innovation. The advantages of instruments that handle this 4-point scale are documented (Chang, 1994), especially in multivariate studies using online surveys. A two-factor model has been reported for the PSS in previous studies with positively worded items representing perceived coping self-efficacy and negative items capturing hopelessness.

*Patient Health Questionnaire-4* (PHQ-4). This instrument assesses depressive and anxiety symptoms (Löwe et al., 2010). This ultra-brief version contains 4 items with a 4-point Likert response format, where 0 corresponds to “not at all” and 3 to “nearly every day”. The scores of PHQ-2 (depression) and GAD-2 (anxiety) range from 0 to 6, with higher scores indicating higher depressive and anxiety symptoms. The Colombian version (Sanabria-Mazo et al., 2023) of PHQ-4 showed adequate internal consistency for depression (Cronbach’s *α* = .79) and anxiety (*α* = .83). This instrument also showed adequate internal consistency in this sample for depression (*α* = .85) and anxiety (*α* = .84).

*Posttraumatic Growth Inventory (PTGI-5).* This instrument assesses posttraumatic growth (Tedeschi & Calhoun, 1996). Specifically, the 5 items (i.e., relationship with others, new possibilities, personal strength, spiritual change, and appreciation for life) with the highest factor loadings were evaluated in a principal component analysis. The 5 items are rated with a 4-point Likert response format, where 0 corresponds to “not at all” and 3 to “nearly every day”. The total score of the PTGI-5 ranges from 0 to 15, with higher scores indicating higher posttraumatic growth. The Colombian version of PTGI-5 (Gómez-Acosta et al., 2023) showed adequate internal consistency (*α* = .79). This instrument showed adequate internal consistency (*α* = .83) in the sample of this study.

*Connor-Davidson Resilience Scale (CD-RISC-2)*. This scale assesses resilience (Connor & Davidson, 2003). This short version contains 2 items with a 4-point Likert response format, where 0 corresponds to “not at all” and 3 to “nearly every day”. The total score of CD-RISC-2 ranges from 0 to 6, with higher scores indicating higher resilience. This instrument showed adequate internal consistency (*α* = .77) in the sample of this study.

### Statistical analysis

A descriptive analysis of sociodemographic variables was conducted using frequencies (*n*) and percentages (*%*), for categorical variables, and means (*M*) and standard deviations (*SDs*), for quantitative variables. Characteristics of the PSS-4 were explored, including item means and standard deviations, skewness and kurtosis, corrected item-total correlations, and between items in each subscale, and between items of different subscales. These correlations were analyzed using the Spearman-Brown correction, considering the brevity of the scales. There were no identified outliers in the analyses and no participants were excluded due to missing data.

Dimensionality of the PSS-4 was tested through confirmatory factor analysis (CFA), using maximum likelihood robust (MLR) as the estimation method. The following factor models were tested: (1) A one-factor model with the four items loading on one latent factor (i.e., psychological stress), (2) a unidimensional model with method effects (correlated error terms on the negative-phrased items), and (3) a two-factor model with two items in each factor (i.e., hopelessness and coping). According to Schermelleh-Engel et al. (2003), Tucker–Lewis’s Index (TLI), Normed Fit Index (NFI), and Comparative Fit Index (CFI) were used to evaluate goodness-off, with >. 90 confidence intervals and the Root Mean Square Error of Approximation (RSMEA) < .08. The invariance (configural, metric, scalar, and strict) of the models was tested by gender, age, income level, and work status in comparable subsamples with random assignment. To determine measurement invariance, the multigroup CFA was conducted, observing a change of ΔCFI that is less than or equal to .01 (Chen, 2007).

To estimate reliability, internal consistency of the PSS-4 was assessed through McDonald’s *ω*, Cronbach’s *α*, and Guttman’s *λ*^2^. Construct validity analysis (convergent and discriminant) was performed using Spearman’s rho statistic considering the nonparametric distribution of the data in this sample. Specifically, psychological stress scores (PSS-4) were correlated with other mental health indicators (measured with ultra-brief instruments), such as resilience (CD-RISC-2) and posttraumatic growth (PTGI-5), as well as anxiety (GAD-2) and depression (PHQ-2). For assessing predictive validity, a hierarchical multiple regression analysis was performed to examine whether perceived stress was a significant predictor of resilience (CD-RISC-2), posttraumatic growth (PTGI-5), anxiety (GAD-2), and depression (PHQ-2). Sociodemographic variables (gender, age, and income level) were entered in the first step, followed by PSS-4 scores were entered in the second step.

Finally, a known-groups validity approach was used to estimate associations between PHQ-4 scores and socio-demographic characteristics (i.e., gender, age, income level, and work status). To this end, univariate group comparisons were performed with the PSS-4 scores as dependent variables through t-tests and one-way analysis of variance (ANOVA). Statistical analyses were performed with SPSS with AMOS29®, JAMOVI 2.3®, and JASP 0.17.1.®.

## Results

### Item analysis

Table 2 shows descriptive analyses of items and total scale (PSS-4). Preliminary analyses showed that the items scores of the PSS-4 were not normally distributed, presenting positive skewness and platykurtic tendency. Subsequently, when carrying out a Kolmogorov-Smirnov analysis, it was confirmed that the distribution of the data was non-parametric. Mean (SD) score of PSS-4 was 3.77 (2.61) and corrected item-total correlation coefficients were all greater than .44 (*p* < .01), thus suggesting good scale homogeneity.

**Table 2.** Characteristics of the items and subscales of the PSS-4.

| **PSS-4 items** | ***M* (95% *CI*)** | ***SD*** | ***S*** | ***K*** | ***r*<sub>tot</sub>** |
| --- | --- | --- | --- | --- | --- |
| 1. In the last month, how often have you felt that you were unable to control important things in your life?<br>*En el último mes, ¿con qué frecuencia ha sentido incapaz de controlar las cosas importantes en su vida?* | 0.86 (0.83 to 0.89) | 0.81 | .83 | .34 | .55 |
| 2. In the last month, how often have you felt confident about your ability to handle your personal problems?<br>*En el último mes, ¿con qué frecuencia ha estado seguro sobre su capacidad para manejar sus problemas personales?* | 1.01 (0.96 to 1.05) | 0.97 | .52 | -.84 | .44 |
| 3. In the last month, how often have you felt that things were going your way?<br>*En el último mes, ¿con qué frecuencia ha sentido que las cosas le van bien?* | 1.05 (1.00 to 1.08) | 0.87 | .45 | -.57 | .59 |
| 4. In the last month, how often have you felt difficulties were piling up so high that you could not overcome them?<br>*En el último mes, ¿con qué frecuencia ha sentido que las dificultades se acumulan tanto que no puede superarlas?* | 0.86 (0.82 to 0.89) | 0.86 | .86 | .13 | .49 |
| Total | 3.77 (3.65 to 3.88) | 2.61 | .40 | -.27 |  |

*Note.* *n* = 1991. PSS-4 = Perceived Stress Scale. *M* = mean; 95% *CI* = 95% confidence interval; *SD* = standard deviation; *S* = Skewness; *K* = Kurtosis. *r*<sub>tot</sub> = corrected item-total correlations. Items scores can range from 0 (“not at all”) to 3 (“nearly every day”).

### Dimensionality

Following a cross-validation approach, the total sample was randomly divided into two subsamples to examine the PSS-4 factor structure. Thus, a principal components analysis was computed with the first subsample and CFAs with the second subsample. The sample size for both the exploratory and confirmatory analyses was adequate, since the recommendation of a minimum of 10 participants per item could be met. Kaiser-Meyer-Olkin (KMO) analyses indicated that the first subsample was significantly sufficient for factor analysis. Consequently, an exploratory analysis (with Promax rotation) was performed, which showed that the items were grouped into a one-factor model that explained 56% of the total variance of the construct. Nevertheless, when conducting the CFA, the PSS-4 was found to have inadequate indicators in the one-factor model. Therefore, a one-factor model was calculated with the second subsample in which all items loaded, but incorporating a correlated residual between negatively phrased items, and a new analysis was performed to compare the two models.

The fit indices for the correlated two-factor model were significantly better (*p* < .001) than one-factor model with the four items loading on one latent factor, but were not significantly better (*p* < .001) than those obtained for the unidimensional model with method effects (correlated error terms on the negative-phrased items). The comparison of the fit indices is displayed in Table 3.

**Table 3.** Confirmatory factor analysis (CFA) comparing fit indices of the one-factor and two-factor model of PSS-4.

| | **CFI** | **TLI** | **NFI** | **RMSEA** | $X^2$ | $X^2/gl$ | $p$ |
| --- | --- | --- | --- | --- | --- | --- | --- |
| Two-factor model | .99 | .98 | .99 | .01 | 0.046 | .046 | .83 |
| Unifactorial model (correlated errors) | .99 | .97 | .99 | .07 | 2.26 | 1.128 | .32 |
| One-factor model | .78 | .35 | .78 | .32 | 424.19 | 212.08 | < .01 |
| Expected | > .90 | > .90 | > .90 | < .10 | ---- | < 3.0 | > .05 |

Regarding factor loadings of the three tested factor models, the one-factor model ranged between .64 and .77, the unidimensional model between .32 to .85, and the two-factor model between .59 to .93 (see Figure 1). Responding to hypothesis 1, these findings confirm that the two-factor model has a significantly better fit to the data than the one-factor model; however, when correlating the error of the negatively phrased items (2 and 3), it is found that they contribute a reduced factor load to the unifactorial model. In this case, the two-factor model shows better global adjustment and factor loading, the reason for which is decided to continue the analysis of invariance with this model.

**Figure 1.**
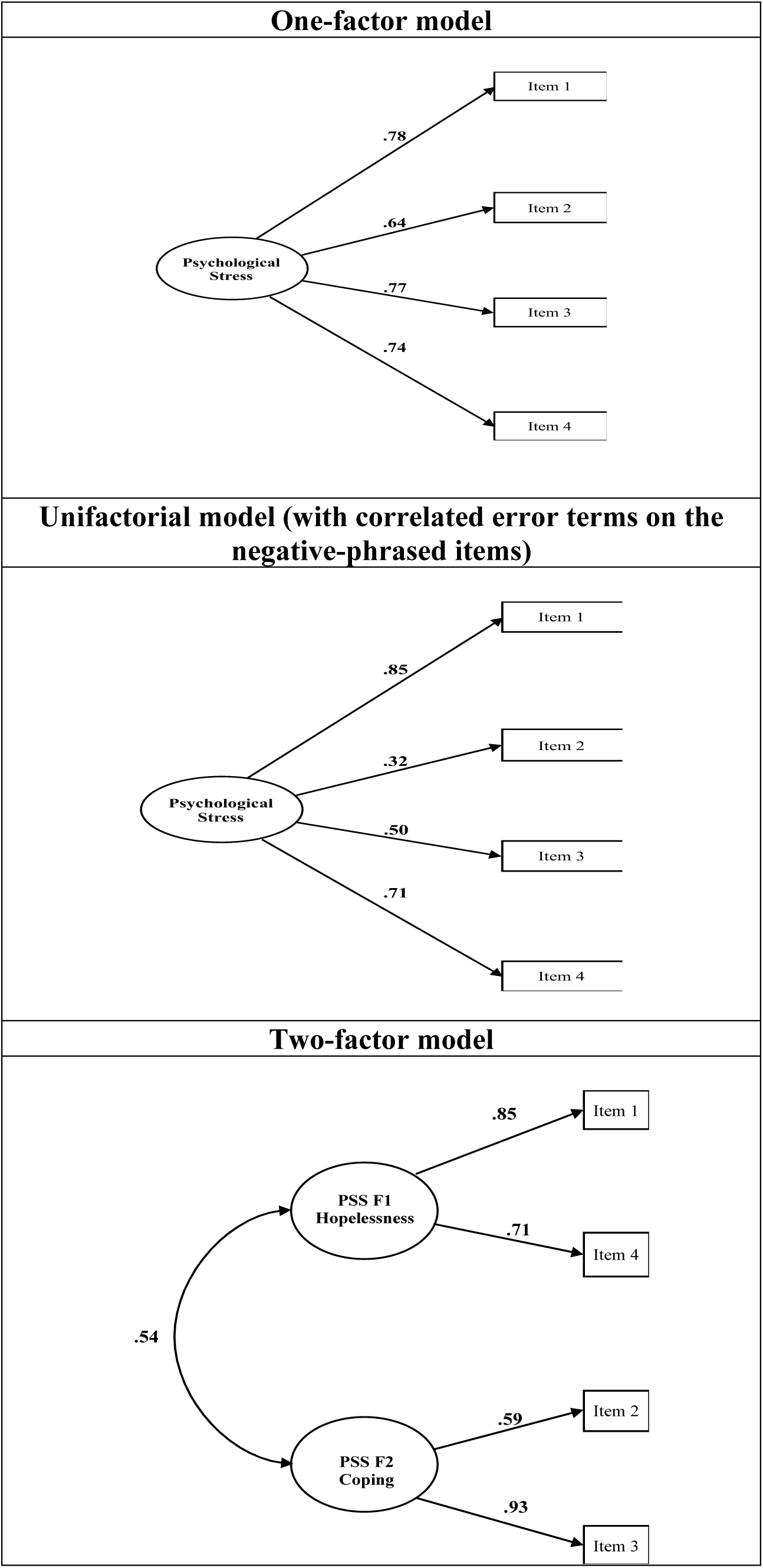
CFA of the PSS-4.

### Configural invariance

As shown in Table 4, comparable subsamples with random assignment were used to evaluate configural invariance by socio-demographic characteristics. No structural differences were found in the best-fitting model according to gender, income level, age, and work status with a Δ CFI < 0.01 and a Δ RMSEA < 0.015. However, this assumption only holds for the configural model (and partially when comparing the metric, scalar, and strict models) for the other analyses, and total factorial invariance of the PSS-4 cannot be concluded concerning the criteria of age, income level, and work status. These results confirm hypothesis 2.

**Table 4.** Test for configurational invariance across gender, age group, income level, education level, and work status using multi-group CFA.

|  | X<sup>2</sup> (df) | ΔX<sup>2</sup> | CMIN/df | CFI | Δ CFI | RMSEA | Δ RMSEA | Invariance |
| --- | --- | --- | --- | --- | --- | --- | --- | --- |
| *Across gender* |  |  |  |  |  |  |  |  |
| Groups |  |  |  |  |  |  |  |  |
| Male (*n* = 402) | 2.018 (1) | --- | 2.018 | .998 | --- | .050 | --- | --- |
| Female (*n* = 444) | 0.048 (1) | --- | 0.048 | 1.00 | --- | .000 | --- | --- |
| Multigroup analysis |  |  |  |  |  |  |  |  |
| Configural model | 2.121 (2) | **.346** | 1.060 | 1.00 | --- | .006 |  | Invariant |
| Metric model | 7.410 (4) | **.116** | 1.102 | .999 | **.001** | .007 | **.001** | Invariant |
| Scalar model | 12.089 (7) | **.098** | 1.727 | .996 | **.003** | .023 | **.014** | Invariant |
| Strict model | 7.535 (4) | **.110** | 1.383 | .990 | **.006** | .026 | **.003** | Invariant |
| *Across age group* |  |  |  |  |  |  |  |  |
| Groups |  |  |  |  |  |  |  |  |
| ≤ 29 years (*n* = 561) | 0.135 (1) | --- | 0.135 | 1.00 | --- | .000 | --- | --- |
| 30 -44 years (*n* = 785) | 0.691 (1) | --- | 0.691 | 1.00 | --- | .000 | --- | --- |
| ≥ 45 years (*n* = 565) | 0.124 (1) | --- | 0.124 | 1.00 | --- | .000 | --- | --- |
| *Multigroup analysis* |  |  |  |  |  |  |  |  |
| Configural model | 3,594 (4) | **.464** | 0.898 | 1.00 | --- | .000 | --- | Invariant |
| Metric model | 175.130 (8) | < .00 | 21.891 | 1.00 | **.000** | .000 | **.000** | Invariant |
| Scalar model | 83.451 (14) | < .00 | 5.960 | .898 | .102 | .080 | .080 | Variant |
| Strict model | 40.049 (8) | < .00 | 5.006 | .875 | .023 | .069 | **.011** | Variant |
| *Across income level* |  |  |  |  |  |  |  |  |
| Groups |  |  |  |  |  |  |  |  |
| Low (*n* = 490) | 7.404 (1) | --- | 7.404 | .958 | --- | .196 | --- | --- |
| Medium (*n* = 1250) | 0.003 (1) | --- | 0.003 | 1.00 | --- | .000 | --- | --- |
| High (*n* = 171) | 0.869 (1) | --- | 0.869 | 1.00 | --- | .000 | --- | --- |
| *Multigroup analysis* |  |  |  |  |  |  |  |  |
| Configural model | 4.309 (4) | **.366** | 1.077 | .989 |  | .060 |  | Invariant |
| Metric model | 83.800 (8) | < .01 | 10.475 | .989 | **.000** | .041 | .019 | Variant |
| Scalar model | 59.376 (14) | < .01 | 4.241 | .835 | .154 | .106 | .065 | Variant |
| Strict model | 35.117 (8) | < .01 | 4.389 | .842 | **.007** | .095 | **.011** | Invariant |
| *Across work status* |  |  |  |  |  |  |  |  |
| Groups |  |  |  |  |  |  |  |  |
| Formal workers (*n* = 336) | 0.995 (1) | --- | 0.995 | 1.00 | --- | .000 | --- |  |
| Formal workers (*n* = 331) | 0.499 (1) | --- | 0.499 | 1.00 | --- | .000 | --- |  |
| Unemployed (*n* = 329) | 3.320 (1) | --- | 3.320 | .993 | --- | .084 | --- |  |
| Other (*n* = 182) | 0.064 (1) | --- | 0.064 | 1.00 | --- | .000 | --- |  |
| *Multigroup analysis* |  |  |  |  |  |  |  |  |
| Configural model | 5.415 (6) | **.492** | 0.902 | .999 |  | .004 |  | Invariant |
| Metric model | 61.042 (12) | < .01 | 5.086 | 1.00 | **.001** | .009 | **.005** | Invariant |
| Scalar model | 26.981 (9) | < .01 | 2.997 | .958 | .042 | .061 | .052 | Variant |
| Strict model | 23.739 (12) | .022 | 1.978 | .933 | .025 | .104 | .041 | Variant |

### Reliability

Internal consistency was adequate for the PSS-4 total scale (*α* =.73, *ω* = .72, *λ^2^*= .74), as well as for its two factors: hopelessness (*α* =.75, *ω* = .75, *λ^2^* = .75) and coping (*α* =.71, *ω* = .71, λ^2^ = .71). See Appendix A in the Supplementary Materials for more information on the variability of these indices with the exclusion of specific items from the PSS-4. The reliability indices of the PSS-4 confirm hypothesis 3.

### Construct validity

As for the convergent and discriminant validity analysis, positive significant correlations of the PSS-4 were identified with depression (*r* = .59) and anxiety (*r* = .55), as well as significant negative with post-traumatic growth (*r* = –.37) and resilience (*r* = –.47). These indicators rule out collinearity between the measures explored (see Table 5). All these associations were significant at the p < .01. These findings are consistent with hypothesis 4.

**Table 5.** Construct validity analysis (convergent and divergent) of PSS-4.

|  | Convergent validity |  | Discriminant validity |  |
| --- | --- | --- | --- | --- |
|  | Depression<br>(PHQ-2) | Anxiety<br>(GAD-2) | Posttraumatic<br>growth (PTGI-5) | Resilience<br>(CD-RISC-2) |
| Total scale (PSS-4) | .59** | .55** | -.37** | -.47** |
| Factor 1 (hopelessness) | .62** | .59** | -.31** | -.42** |
| Factor 2 (coping) | .43** | .39** | -.34** | -.42** |

*Note.* \*\* Correlation is significant at the 0.01 level (2-tailed).

### Predictive validity

As shown in Table 6, hierarchical multiple regression analysis showed that elevated PSS-4 scores significantly (*p* <.001) predicted depression (R^2^ = .39, Beta = 0.38) and anxiety scores (R^2^ = .34, Beta = 0.37). Likewise, PSS-4 low scores significantly (*p* <.001) predicted resilience (R^2^ = .24, Beta = –0.21) and post-traumatic growth (R^2^ = .15, Beta = –0.47). Although the sociodemographic variables contribute to the prediction in the first step of the model, their contribution to the explained variance was not significant, in contrast to the PSS-4 scores to the explanation of the psychological variables assumed as dependent. These results confirm hypothesis 5.

**Table 6.** Predictive validity of the PSS-4.

| **Predict variable** | **F (*p* value)** | **R<sup>2</sup>** | **Beta (CI 95%)** |
| --- | --- | --- | --- |
| Depression (PHQ-2) | 1222.35 (<.001) | .39 | 0.38 (-0.40 to 0.36) |
| Anxiety (GAD-2) | 964.93 (<.001) | .34 | 0.37 (-0.39 to 0.35) |
| Resilience (CD-RISC-2) | 607.81 (<.001) | .24 | -0.21 (-0.23 to -0.29) |
| Posttraumatic growth (PTGI-5) | 330.98 (<.001) | .15 | -0.47 (-0.53 to -0.42) |

### Known-groups validity

A comparative analysis of the PSS-4 was carried out by socio-demographic characteristics. As shown in Table 7, statistically significant differences were found according to age, income level, and work status, but not according to gender. Significantly higher scores on the PSS-4 were observed among young people, as well as respondents with lower incomes and those unemployed. Contrary to hypothesis 5, young and unemployed people scored higher on the PSS-4 than older and employed people. People with lower levels of income scored higher on the PSS-4, being consistent with hypothesis 6.

**Table 7.** Association PSS-4 scores and socio-demographic characteristics.

| **Variables** | **Score**<br>***M (SD)*** | **Difference groups**<br>**(*p* value)** |
| --- | --- | --- |
| *Gender* |  | .07 |
| 1. Male | 3.86 (2.61) | 1 > 2 |
| 2. Female | 3.54 (2.61) |  |
| *Age group* |  | < .001 |
| 1. 18-29 years | 4.65 (2.69) | 1 > 2 > 3 |
| 2. 30-44 years | 3.64 (2.51) |  |
| 3. ≥ 45 years | 2.82 (2.34) |  |
| *Income level* |  | < .001 |
| 1. Low | 4.81 (2.71) | 1 > 2 > 3 |
| 2. Medium | 3.52 (2.45) |  |
| 3. High | 2.60 (2.56) |  |
| *Work status* |  | < .001 |
| 1. Employed formal | 3.50 (2.50) | 3 < 1, 2, and 4 |
| 2. Employed informal | 3.65 (2.51) |  |
| 3. Unemployed | 4.82 (2.70) |  |
| 4. Others | 3.19 (2.80) |  |

*Note.* Between-group differences were calculated through t-test and ANOVA

## Discussion

The purpose of the current study was to evaluate the psychometric properties of the PSS-4 in a large sample of Colombians with university education. The first objective was to examine the factorial structure. Unlike the Ecuadorian version of Ruisoto et al. (2020), the CFA showed better fit indices for the two-factor model (hopelessness and coping) in this sample, considering that the two-factor version obtained more homogeneous variance weights contribute by the items concerning the structure arranged in previous two-factor versions of PSS-10 (Campo-Arias et al., 2021; Perera et al., 2017) and PSS-4 (Boluarte-Carbajal et al., 2023), as well as optimal fit indicators in all parameters (Schermelleh-Engel et al., 2003). These findings confirm hypothesis 1. In fact, the factorial distribution of the PSS-4 items into one factor for hopelessness (item 1 and 4) and another for coping (item 2 and 3) is consistent with the documented theoretical relationship between the two constructs (Chirico et al., 2017; Morris et al., 2018).

Although in this sample the two-factor model obtained the best-fit indices, this may be because the two-factor structure obtained in previous studies was probably an artifact of the response styles associated with item wording (Hankins, 2008), discussion inherent in the field of psychometrics (Aguado et al., 2012; Chyung et al., 2018; Dalal & Carter, 2015). However, both models presented adequate psychometric properties, supporting the argument that the one-factor model might also be suitable for the implementation in the clinical setting, as has been suggested (Ruisoto et al., 2020).

The second objective was to investigate invariance (configural, metric, scalar, and strict) of the best-fitting model across socio-demographic characteristics. The PSS-4 was invariant in the configural model according to gender, age, income level, and work status. However, in the multigroup analyses, invariance was only detected in all models (configural, metric, scalar, and strict) by gender, being consistent with previous validations of PSS-10 (Barbosa-Leiker et al., 2013; Lee, 2022; Liu et al., 2020; Reis et al., 2019). This could be because the sample of this study presented some differences in age, income level, and work status, which could indirectly affect the factorial invariance analysis (Chen, 2007). The two-factor model was invariant by gender in this sample supporting hypothesis 2.

The third and fourth objectives were to determine the reliability and construct validity of the PSS-4, respectively. Internal consistency was acceptable (above .70), with results like those reported by Ruisoto et al. (2020) in Ecuador for the PSS-4, but lower than those achieved in Colombia for the PSS-14 and PSS-10 before (Campo-Arias et al., 2009, 2014) and after the pandemic (Campo-Arias et al., 2021) in samples of participants with university education. It’s well known that Cronbach’s alpha is influenced by the number of items in the tested scale. For shorter tests, it has naturally reduced values (Tavakol & Dennick, 2011). Consistent with previously available evidence, the convergent validity between psychological stress scores with anxiety and depression (Perera et al., 2017), as well as the discriminant validity with posttraumatic growth (Gómez-Acosta et al., 2023) and resilience (Ruisoto et al., 2020) was in the expected direction. These findings are consistent with hypothesis 3 and 4.

The fifth objective was to evaluate the predictive validity of the PSS-4 on a set of psychological variables (resilience, posttraumatic growth, anxiety, and depression,). The PSS-4 has an adequate capacity to predict possible depressive and anxiety symptoms (APA, 2023; Feng et al., 2023), as well as acceptable levels of prediction of protective factors such as resilience (Finstad et al., 2021) and post-traumatic growth (Gómez-Acosta et al., 2023), confirming hypothesis 5. Accordingly, the ultra-brief version of the PSS-4 could be used as an input to obtain possible indicators of mental health impairment and for the development of primary care actions, particularly in Colombian professionals exposed to multiple psychosocial stressors in the different work environments where they work (Stringer, 2023a), such as precarious employment contracts, long working hours, lack of social benefits and performance of their duties in dangerous contexts, among others (Gómez-Acosta et al., 2022).

Finally, the sixth objective was to test the relationship between PPS-4 scores and sociodemographic characteristics of this sample. This study provides further evidence about gender, age, income level, and work status as risk factors for psychological stress. Consistent with other studies and with hypothesis 6, it was observed that people with higher educational level presented lower scores in psychological stress (Vallejo et al., 2018; Warttig et al., 2013; Ruisoto et al., 2020). Contrary to what was hypothesized, it was observed that younger and unemployed individuals reported higher scores than older and employed individuals. These findings could be explained considering the large amount of empirical evidence reflecting the negative effect generated by the COVID-19 pandemic on mental health in the short (Hossain et al., 2020) and medium-long term (Newnham et al., 2022).

## Limitations

The following limitations should be kept in mind when interpreting these results. The first limitation is that the analyzes were performed on a non-representative sample, which prevents the generalization of these results to the population with university education. The second limitation is that the sampling (aimed exclusively at adults with university education) limited the representation of people with other sociodemographic characteristics. The third limitation is that temporal stability was not calculated in this study, which could help to explore the performance of this instrument over time. The fourth limitation is related to the impossibility of comparing the psychological stress scores obtained with the self-report with biomarkers (i.e., cortisol) under controlled conditions so that the scale sets the optimal cut-off point (considering the indicators of specificity and sensitivity). Finally, the fifth limitation is that online data collection may negatively affect the representation of population groups with difficulties connecting to the Internet (self-selection bias).

## Conclusions

In summary, the current findings have showed that the PSS-4 is a reliable and valid ultra-brief self-administered instrument for measuring psychological stress in the Colombian population with university education. The CFA showed significantly adequate fit indices for the two-factor model (hopelessness and coping), being invariant across gender, income level, age, and work status. Internal consistency was adequate for the PSS-4. The convergent and discriminant validity, as well as the predictive validity, explored was consistent with the theoretically expected direction. Significantly higher scores on the PSS-4 were observed among young people, as well as among people with lower incomes and the unemployed. Even though in this sample the two-factor model obtained the best fit indices, the one-factor model could also be suitable for implementation in the clinical setting. For this reason, the PSS-4 can be useful as a screening tool for psychological stress in clinical and non-clinical populations, especially as a complement to the assessment and monitoring processes, considering its adequate psychometric properties. Finally, PSS-4 is a brief screening tool that can help optimize the time resources of health systems or reduce the burden of questionnaires in research that aim to assess various constructs.

## Data Availability

All data produced are available online at https://doi.org/10.6084/m9.figshare.21701231

## Supplementary Material

## Appendix A

Internal consistency analysis of the PSS-4.

| Internal consistency |  |  |  |
| --- | --- | --- | --- |
| Total scale (PSS-4) | $\omega$ (IC 95%) | $\alpha$ (IC 95%) | $\lambda^2$ (IC 95%) |
|  | 0.72 (0.70-0.74) | 0.73 (0.71-0.75) | 0.74 (0.71-0.76) |
| If item dropped |  |  |  |
| Item | $\omega$ | $\alpha$ | $\lambda^2$ |
| Item 1 | 0.69 | 0.65 | 0.66 |
| Item 2 | 0.73 | 0.72 | 0.72 |
| Item 3 | 0.65 | 0.62 | 0.63 |
| Item 4 | 0.71 | 0.68 | 0.69 |
| Factor 1 (hopelessness) | 0.75 | 0.75 | 0.75 |
| Factor 2 (coping) | 0.71 | 0.71 | 0.71 |

